# The Prevalence of Tuberculosis Infection Among Foreign-Born Canadians: A Modelling Study

**DOI:** 10.1101/2023.03.01.23286631

**Authors:** Aria Ed Jordan, Ntwali Placide Nsengiyumva, Rein M. G. J. Houben, Peter J. Dodd, Katie D. Dale, James M. Trauer, Justin T. Denholm, James C. Johnston, Faiz Ahmad Khan, Jonathon R. Campbell, Kevin Schwartzman

## Abstract

**Background:** The prevalence of tuberculosis infection (TBI) is critical to designing tuberculosis (TB) prevention strategies, yet it is unknown in Canada. We estimated TBI prevalence among foreign-born Canadians.

**Methods:** Using a previously developed Gaussian process regression model, annual risk of infection trends abroad were constructed and used to estimate TBI prevalence by age and year of migration to Canada for persons from each of 168 countries. These stratified TBI prevalence estimates were combined with Canadian census data to estimate overall TBI prevalence among foreign-born residents during census years 2001, 2006, 2011, and 2016. We also estimated TBI prevalence according to age, WHO-estimated TB incidence in country of origin, and province/territory of residence. Finally, we estimated the prevalence of TBI acquired within the two preceding years.

**Results:** Estimated TBI prevalence among foreign-born Canadians overall was 25% (95% uncertainty interval [UI]: 20-35%), 24% (20-33%), 23% (19-30%), and 22% (19-28%) for census years 2001, 2006, 2011, and 2016, respectively. TBI prevalence increased with age at migration and TB incidence in the country of origin. In 2016, estimated TBI prevalence was lowest in Quebec (19%, 95% UI: 16-25) and highest in Alberta and British Columbia, at 24% each. Among all foreign-born Canadian residents with TBI in 2016, we estimated that only 1 in 440 were infected within the two preceding years.

**Interpretation:** Approximately 1 in 4 foreign-born Canadians has TBI; estimated prevalence has remained quite stable over the last two decades. However, a very small minority of people with TBI were infected within the last two years—the highest risk period for progression to TB disease. These data may inform future TBI screening policies.

## INTRODUCTION

Before the COVID-19 pandemic, tuberculosis (TB) disease was the leading cause of death by a single infectious agent worldwide, and it remains the second leading cause. TB disease is preceded by TB infection (TBI), which traditionally has been identified by testing for immunoreactivity to the causative agent, *Mycobacterium tuberculosis*. An estimated 23% of the world’s population has TB infection (TBI).^1^ Depending on sociodemographic and medical characteristics, the World Health Organization (WHO) suggests that 5-10% of persons with TBI will progress to TB disease, the cause of 1.6 million deaths in 2021.^2,3^

The WHO has established targets for TB disease elimination in low-incidence countries such as Canada, with a goal to reduce annual TB disease incidence to 1 case per million population. TB disease incidence in Canada declined steadily through the second half of the 20^th^ century but has remained essentially unchanged over the last decade. Despite a previous target of reducing TB incidence to 3.5 per 100,000 by 2015, TB incidence in Canada remained 4.7 per 100,000 in 2020. ^4–6^ The lack of further progress toward elimination is the result of epidemiologic shifts without the implementation of new prevention strategies

In Canada, TB disease disproportionately impacts foreign-born persons, who bear 80% of the TB disease burden, despite representing only 23% of the population.^4,5,7^ TB disease among foreign-born persons is largely due to progression of TBI acquired within their countries of origin.^4^ Tuberculosis preventive treatment (TPT) is highly effective at preventing progression to TB disease. However, implementation of TPT in Canada is limited.^8,9^ With available TBI diagnostics, widespread TBI screening and treatment programs would likely be cost-prohibitive^10^ and could result in the provision of treatment to large numbers of people who may derive minimal benefit.^11,12^ Instead, targeting screening to those at highest risk of TBI would improve cost-effectiveness as well as health gains for treated individuals.^13^ However, TBI prevalence among foreign-born Canadians is unknown. To support the design and implementation of TB prevention programs, we estimated TBI prevalence and patterns among foreign-born Canadians.

## METHODS

We used estimates of the annual risk of TBI—the probability someone would become infected with *M. tuberculosis* each year—from 168 countries between 1889 and 2016 to estimate the probability of TBI among people immigrating to Canada according to year of birth, age at arrival in Canada, and country of origin. We integrated Canadian census data from 2001, 2006, 2011, and 2016 to estimate the prevalence of TBI among foreign-born residents of Canada during each census year.

### Data Sources

#### Annual Risk of Infection Data

We adapted a previously published approach to obtain country-specific estimates for the annual risk of TBI. In brief, estimated historic annual risks of infection (ARI) were constructed using two data sources. In-country tuberculin skin test (TST) surveys were used to directly estimate ARI where available (37 countries).^1,14^ When TST surveys were unavailable, historic ARIs were estimated based on World Health Organization (WHO) estimates of the prevalence of smear-positive TB and the documented relationship between smear-positive pulmonary TB prevalence and annual infection risk,^15–17^ while accounting for the impact of HIV infection in each country.^1^ The delta method was used to propagate uncertainty and estimate variance around ARI estimates.^18^ Countries were excluded from analysis if their population was below 500,000 people, if they could not be matched with TB disease incidence data, or if neither TST survey data nor adequate TB disease prevalence data were available. A total of 50 countries were excluded. The total number of people living in Canada from these 50 countries combined ranged from 66,670 (1.3% of all foreign-born residents) to 83,180 (1.1%) between the 2001 and 2016 census years, respectively (Table S1).^1^ This left 168 countries of origin in our analysis.

#### Census Data

Canadian census data detailing the number of permanent residents and foreign-born Canadian citizens living in Canada (hereafter collectively referred to as foreign-born residents) were obtained from Statistics Canada for years 2001, 2006, 2011 and 2016.^19^ These data include the number of permanent residents and citizens from each country (Table S2), further grouped into categories based on age and year of immigration (Table S3). Within each age and year of immigration category, we assumed a uniform distribution of people for each specific age and year of immigration. Temporary workers, visitors, and students were excluded.

### Definitions

Annual TB disease incidence in country of origin per 100,000 person-years was categorized into groups of 0-9, 10-49, 50-99, 100-199, and ≥200 based on WHO global TB disease incidence estimates from 2016.^20,21^ TB disease incidence estimates from 2016 were used as this was the final census year analyzed, and TB disease incidence data prior to year 2000 are considered unreliable.^22^ Year of birth was calculated as the census year minus reported age. Year of immigration was considered the year in which individuals acquired permanent residence. Age at immigration was calculated as year of immigration minus year of birth. We categorized age during each census year and age at immigration as: <15 years, 15-34 years, 35-54 years, 55-74 years, ≥75 years.

### Data Analysis

To account for uncertainty around ARI estimates, a Gaussian process regression model was used to simulate 200 log-scale ARI trends for each country and year from 1934 to 2014, conditioned on the available data. The ARI estimate for 2014 was used for the years 2015 and 2016, while the ARI estimate for 1934 was used for previous years back to 1889, giving ARI estimates based on country and year from 1889-2016.

For each of the 200 sampled log-scale ARI trends, probabilities of TBI for each combination of country, birth year, and migration year were generated as follows (see Appendix Figure S1 for further details). First, we calculated annual risks of TBI, assuming risks were uniform among people from the same country, birth year, and migration year, by exponentiating each log-ARI. Second, we summed annual risks of infection for each country, birth year, and migration year to estimate the cumulative risk of infection for each census year (i.e., to 2001, 2006, 2011, and 2016)—accounting for annual risks of infection in country of origin before migration, as well as within Canada after migration. Since precise calendar dates of migration were unknown, we assumed that the average date of arrival for each cohort was the mid-point of the calendar year, resulting in the risk of infection during the year of migration being an average of the risk of infection in the country of origin and in Canada. Finally, using the cumulative risk of infection, we calculated the probability of TB infection for each combination of birth year, country of origin, and migration year.

For each census year, by summing relevant strata, we estimated the number of foreign-born residents living in Canada overall and by subgroups. These subgroups reflected categorization by age at immigration, age during each census year, TB disease incidence per 100,000 persons in country of origin, and the most frequent countries of origin across census years. Using our estimates of the probability of TBI, we estimated the prevalence of TBI for each census year overall, for each of these subgroups, and by time since immigration. Finally, for the 2016 census year only, we estimated the prevalence of TBI in Ontario, Quebec, Alberta, and British Columbia (the four largest immigrant-receiving provinces), and the rest of Canada, and estimated the prevalence of TBI acquired within the last 2 years. The point estimate and 95% uncertainty intervals (95% UI) for TBI estimates were calculated as the 50^th^, 2.5^th^, and 97.5^th^ percentile, respectively. Differences in median TBI prevalence across census years were tested using the Kruskal-Wallis test. All analyses were performed using R (version 4.0.2).

### Ethics Approval

This study involved the use of publicly available, de-identified, aggregate data. Approval by a research ethics board was not required.

## RESULTS

### POPULATION COMPOSITION FROM 2001 TO 2016

During the 2016 census year, 7.5 million foreign-born residents lived in Canada. This represented an increase of 2.1 million (38%) since 2001. The median age (IQR) of foreign-born residents in each census year from 2001 to 2016 was 46 (32-58), 47 (32-61), 47 (32-61), and 48 (33-62) years, respectively; 74% had immigrated before the age of 35. In each of the census years, 52% of foreign-born residents were women while 48% were men. Between the 2001 and 2016 census years, the number of people immigrating from Europe decreased in absolute number and proportion, while those from Asia increased in both absolute number and proportion. In line with these trends, the proportion of foreign-born residents in Canada from countries with TB disease incidence ≥200 per 100,000 per year increased from 15% to 25% between 2001 and 2016, while the proportion from countries with TB incidence <10 per 100,000 decreased from 38% to 25% during the same period (Table 2).

**Table 1.** Composition of the foreign-born Canadian population in census years 2001-2016.

| Population | Census Year |  |  |  |
| --- | --- | --- | --- | --- |
|  | 2001 | 2006 | 2011 | 2016 |
| **Total Foreign-Born Residents** | 5,448,480 | 6,524,190 | 7,217,295 | 7,539,895 |
| **Sex, N (%)** |  |  |  |  |
| Female | 2,825,870 (52) | 3,389,085 (52) | 3,763,070 (52) | 3,953,825 (52) |
| Male | 2,622,610 (48) | 3,135,105 (48) | 3,454,225 (48) | 3,586,070 (48) |
| **Age during census year** |  |  |  |  |
| Median (IQR) age, years | 46 (32-58) | 47 (32-61) | 47 (32-61) | 48 (33-62) |
| **Age during census year, N (%)** |  |  |  |  |
| <15 years | 301,470 (6) | 336,705 (6) | 372,190 (6) | 373,960 (5) |
| 15-34 years | 1,207,909 (23) | 1,369,695 (23) | 1,493,443 (23) | 1,608,233 (22) |
| 35-54 years | 2,011,356 (38) | 2,184,035 (36) | 2,400,517 (37) | 2,642,352 (35) |
| 55-74 years | 1,189,814 (23) | 1,478,539 (25) | 1,519,770 (23) | 1,963,148 (26) |
| ≥75 years | 565,341 (11) | 666,871 (11) | 706,630 (11) | 877,517 (12) |
| **Region of Origin, N (%)** |  |  |  |  |
| Americas | 834,260 (15) | 946,140 (15) | 1,058,010 (16) | 1,127,815 (15) |
| Europe | 2,282,795 (42) | 2,267,440 (37) | 2,127,790 (31) | 2,088,405 (28) |
| Africa | 281,445 (5) | 373,340 (6) | 492,025 (7) | 637,485 (8) |
| Asia | 1,986,105 (37) | 2,521,035 (41) | 3,041,100 (45) | 3,629,165 (48) |
| Oceania | 47,825 (0.9) | 52,625 (0.9) | 54,530 (0.8) | 56,920 (0.8) |
| **TB Disease Incidence in Country of Origin in 2016, N (%)** |  |  |  |  |
| 0-9 per 100,000 persons | 2,017,280 (38) | 2,019,465 (33) | 1,884,040 (29) | 1,890,965 (25) |
| 10-49 per 100,000 persons | 1,011,810 (19) | 1,144,505 (19) | 1,257,380 (19) | 1,480,190 (20) |
| 50-99 per 100,000 persons | 1,098,005 (21) | 1,341,575 (22) | 1,450,320 (22) | 1,671,545 (22) |
| 100-199 per 100,000 persons | 332,735 (6) | 411,935 (7) | 473,735 (7) | 579,945 (8) |
| ≥200 per 100,000 persons | 816,060 (15) | 1,118,365 (19) | 1,427,075 (22) | 1,842,565 (25) |
| **Age at Immigration, N (%)** |  |  |  |  |
| <15 years | 1,613,358 (31) | 1,828,028 (30) | 2,253,488 (35) | 2,504,326 (34) |
| 15-34 years | 2,291,796 (43) | 2,655,510 (44) | 2,626,257 (41) | 3,007,347 (40) |
| 35-54 years | 1,140,530 (22) | 1,303,154 (22) | 1,349,278 (21) | 1,643,312 (22) |
| 55-74 years | 205,982 (4) | 225,683 (4) | 240,426 (4) | 279,984 (4) |
| ≥75 years | 24,224 (0.5) | 23,470 (0.4) | 23,101 (0.4) | 30,241 (0.4) |

**Table 2.** Number and proportion of foreign-born Canadians from top ten countries of origin in each census year

| Census Year |  |  |  |  |  |  |  |
| --- | --- | --- | --- | --- | --- | --- | --- |
| 2001 |  | 2006 |  | 2011 |  | 2016 |  |
| Place of Origin | N (%) | Place of Origin | N (%) | Place of Origin | N (%) | Place of Origin | N (%) |
| India | 314,265 (11) | India | 443,120 (15) | India | 538,325 (17) | India | 668,565 (18) |
| China | 332,235 (12) | China | 465,940 (15) | China | 533,035 (17) | China | 649,260 (18) |
| Philippines | 232,330 (8) | Philippines | 302,860 (10) | Philippines | 440,925 (14) | Philippines | 588,310 (16) |
| United Kingdom | 604,380 (22) | United Kingdom | 574,740 (19) | United Kingdom | 525,475 (16) | United Kingdom | 499,125 (14) |
| United States of America | 236,225 (9) | United States of America | 247,870 (8) | United States of America | 256,970 (8) | United States of America | 253,700 (7) |
| Italy | 315,215 (11) | Italy | 296,250 (10) | Italy | 255,350 (8) | Italy | 236,635 (7) |
| Hong Kong | 235,315 (8) | Hong Kong | 215,080 (7) | Hong Kong | 191,405 (6) | Hong Kong | 208,945 (6) |
| Poland | 180,150 (6) | Poland | 169,740 (6) | Pakistan | 155,330 (5) | Pakistan | 202,250 (6) |
| Portugal | 153,445 (6) | Viet Nam | 159,940 (5) | Viet Nam | 154,665 (5) | Viet Nam | 169,265 (5) |
| Germany | 173,515 (6) | Germany | 170,165 (6) | Germany | 150,085 (5) | Iran | 154,430 (4) |
| **Total** | **2,777,075** | **Total** | **3,045,705** | **Total** | **3,201,565** | **Total** | **3,630,485** |

From 2001 through 2016 census years, seven countries/jurisdictions (China, India, Italy, Philippines, the United Kingdom, the United States of America, and Hong Kong) were consistently among the ten most frequent places of origin for foreign-born residents. All except the United States of America, Italy, and the United Kingdom have TB disease incidence ≥50 per 100,000 persons, while TB disease incidence is ≥200 per 100,000 persons in India and the Philippines. In the 2016 census year, Pakistan, Viet Nam, and Iran also ranked among the ten most common countries of origin (Table 3); all three have TB disease incidence >100 per 100,000 persons.

**Table 3.** Estimated tuberculosis infection prevalence among foreign-born residents from the seven most frequent countries/jurisdictions of origin

| Place of Origin | TB Infection Prevalence, % (95% UI) |  |  |  | Comparison, 2016 vs. 2001* |
| --- | --- | --- | --- | --- | --- |
|  | Census 2001 | Census 2006 | Census 2011 | Census 2016 |  |
| India | 34 (22-47) | 33 (23-45) | 31 (23-41) | 30 (23-39) | P<0.0001 |
| China | 46 (14-73) | 38 (13-63) | 31 (12-53) | 28 (12-48) | P<0.0001 |
| Philippines | 54 (22-79) | 51 (24-74) | 47 (27-65) | 45 (29-61) | P<0.0001 |
| United Kingdom | 3 (1-40) | 3 (1-36) | 3 (1-30) | 3 (1-26) | P=0.565 |
| United States of America | 10 (1-54) | 8 (1-47) | 6 (1-37) | 5 (1-35) | P< 0.0001 |
| Italy | 5 (1-58) | 5 (1-57) | 5 (1-54) | 4 (1-45) | P=0.800 |
| Hong Kong | 20 (4-65) | 19 (4-64) | 19 (4-62) | 19 (5-65) | P=0.940 |

*UI = uncertainty interval*
*\*P values from Kruskal-Wallis test*

### ESTIMATED TBI PREVALENCE

The overall estimated TBI prevalence among foreign-born residents did not change substantially over time, with a prevalence of 25% (95% UI: 13-35%), 24% (95% UI: 14-33%), 23% (95% UI: 14-30%), and 22% (95% UI: 20-28%) in census years 2001, 2006, 2011, and 2016, respectively.

Between 2001 and 2016, estimated TBI prevalence decreased among foreign-born residents from India, China, Philippines, and the United States, (p < 0.0001 for each, Kruskal-Wallis test)— most substantially for China (Table 4).

**Table 4.** Estimated tuberculosis infection prevalence (95% uncertainty interval) among foreign-born persons by age at immigration and 2016 TB disease incidence per 100,000 persons in country of origin.

| **TB Incidence in Country of Origin** | **Age at Immigration <15** | **Age at Immigration 15-34** | **Age at Immigration 35-54** | **Age at Immigration 55-74** | **Age at Immigration ≥75** | **All** |
| --- | --- | --- | --- | --- | --- | --- |
| **0-9 per 100,000 persons** | 6 (2-21) | 11 (5-22) | 13 (6-23) | 17 (8-35) | 22 (9-47) | 9 (5-22) |
| **10-49 per 100,000 persons** | 7 (4-15) | 14 (9-24) | 22 (15-31) | 34 (23-52) | 39 (24-61) | 14 (9-22) |
| **50-99 per 100,000 persons** | 7 (4-13) | 21 (14-29) | 39 (25-51) | 60 (34-79) | 72 (38-90) | 24 (16-33) |
| **100-199 per 100,000 persons** | 14 (7-24) | 36 (23-50) | 53 (38-66) | 75 (50-89) | 78 (58-93) | 34 (22-46) |
| **≥200 per 100,000 persons** | 12 (9-15) | 35 (29-42) | 54 (44-64) | 67 (54-83) | 75 (62-90) | 35 (22-46) |
| **All** | 8 (6-16) | 22 (19-26) | 37 (31-43) | 55 (45-63) | 65 (50-74) | 22 (19-28) |

In 2016, estimated TBI prevalence varied from 9% (95% UI: 5-22) among persons from countries with TB disease incidence of <10 per 100,000 persons to 35% (95% UI: 22-46) among persons from countries with incidence ≥200 per 100,000 persons. Similarly, it increased markedly with age at immigration, ranging from 8% (95% UI: 6-16) for those who had arrived at <15 years of age to 65% (95% UI: 50-74) for those who had arrived at ≥75 years of age (Table 5). Findings were generally similar in earlier census years (Appendix Table S4).

**Table 5.** Estimated tuberculosis infection prevalence by age at immigration and province/territory of residence in 2016.

| TB Infection Prevalence, % [95% UI] |  |  |  |  |  |  |
| --- | --- | --- | --- | --- | --- | --- |
|  | Age at Immigration |  |  |  |  |  |
| Province/Territory of Residence in 2016 | 0-14 | 15-34 | 35-54 | 55-74 | ≥75 | All |
| Alberta | 8 (6-14) | 24 (21-28) | 40 (35-46) | 58 (49-67) | 69 (56-79) | 24 (21-29) |
| British Columbia | 9 (7-18) | 23 (19-29) | 38 (32-48) | 58 (45-71) | 68 (50-79) | 24 (20-30) |
| Ontario | 8 (6-16) | 22 (18-26) | 36 (30-43) | 53 (43-64) | 63 (48-73) | 22 (18-27) |
| Quebec | 5 (5-15) | 20 (16-25) | 31 (27-38) | 50 (42-61) | 61 (48-72) | 19 (16-25) |
| Other | 7 (6-9) | 25 (21-31) | 36 (30-43) | 29 (22-40) | 35 (24-52) | 22 (18-26) |
| **All** | 8 (6-16) | 22 (19-26) | 37 (31-43) | 55 (45-63) | 65 (50-74) | 22 (19-28) |

*UI = Uncertainty Interval*

In 2016, 94% of all foreign-born residents resided in Ontario, Quebec, Alberta, or British Columbia (Appendix Table S5). In these four provinces, estimated TBI prevalence increased with age at immigration, while for the remaining provinces and territories, prevalence appeared similar among all those aged ≥15 years at immigration (Table 5). The estimated prevalence of TBI was lowest in Quebec (19%, 95% UI: 16-25) and highest in Alberta and British Columbia, at 24% (95% UI: 21-29%) and 24% (95% UI: 20-30%), respectively.

Estimated TBI prevalence increased with age during the 2016 census year from 2% (95% UI: 2-2%) among those aged <15 years in 2016 to 37% (95% UI: 27-54%) among those aged ≥75 years in 2016. Overall prevalence did not change substantially by time since arrival. However, when stratified by age during the census year, estimated prevalence decreased by time since arrival, reflecting lower values among those who entered Canada at younger age (Appendix Table S6).

As expected, the estimated prevalence of TBI acquired in the past two years increased with TB disease incidence in country of origin (Table 6). Overall, only an estimated 0.05% (95% UI: 0.04-0.07%) of foreign-born residents in 2016 had acquired TBI in the past two years. This increased to 0.6% (95% UI: 0.5-0.8%) when considering only those who immigrated in 2015 and 2016. These data suggest that among all foreign-born residents with TBI, an estimated one in 440 had been infected within the last two years. For those with TBI who had arrived within the preceding two years, an estimated one in 39 had been infected within the last two years. To detect one person infected within the last two years among all foreign-born residents, 2,000 (95% UI: 1,429-2,500) would need to be tested. To detect one person infected within the last two years among only those foreign-born residents who had arrived in the last two years, 167 (95% CI: 125-200) would need to be tested.

**Table 6.** Estimated prevalence of recently acquired tuberculosis infection among foreign-born residents.

| TB Incidence in Country of Origin per 100,000 persons | All Foreign-Born Residents |  |  | Only Foreign-Born Residents Immigrating from 2015-2016 |  |  |
| --- | --- | --- | --- | --- | --- | --- |
|  | Prevalence of TBI Acquired in Past 2 Years (% , 95% UI) | Prevalence of TBI Overall (% , 95% UI) | Number of TB Infections to identify to detect one person infected in last 2 years* | Prevalence of TBI Acquired in Past 2 Years (% , 95% UI) | Prevalence of TBI Overall (% , 95% UI) | Number of TB Infections to identify to detect one person infected in last 2 years |
| **0-9** | 0.02 (0.01 - 0.05) | 9 (5-22) | 450 | 0.04 (0.03 - 0.06) | 3 (2-5) | 75 |
| **10-49** | 0.03 (0.02 - 0.05) | 14 (9-22) | 467 | 0.1 (0.1 - 0.2) | 8 (6-11) | 80 |
| **50-99** | 0.04 (0.03 - 0.06) | 24 (16-33) | 600 | 0.3 (0.3 - 0.5) | 22 (16-29) | 74 |
| **100-199** | 0.07 (0.05 - 0.09) | 34 (22-46) | 486 | 0.7 (0.6 - 0.9) | 29 (25-35) | 42 |
| **≥200** | 0.12 (0.10 - 0.17) | 35 (22-46) | 292 | 1.2 (0.9 - 1.7) | 37 (32-41) | 31 |
| **All** | 0.05 (0.04 – 0.07) | 22 (19-28) | 440 | 0.6 (0.5 – 0.8) | 23 (21-25) | 39 |

*UI = uncertainty interval*
*\*Number of TB infections that must be diagnosed, to detect one person infected in the last 2 years = Prevalence of TBI overall/Prevalence of TBI acquired in past 2 years*

## INTERPRETATION

Our estimates suggest that almost one quarter of foreign-born residents in Canada have TBI, a proportion which did not substantially change between 2001 and 2016. However, despite the large number of people with TBI, only a small minority were likely to have been infected within the preceding two years—the highest risk period for progression to TB disease.^23^

Our TBI prevalence estimates are concordant with those from other low-incidence settings, notably an analysis that used similar methods to estimate TBI prevalence among migrants living in Australia.^24^ They are also concordant with cross-sectional surveys. Using tuberculin skin tests, the U.S. National Health and Nutrition Examination Survey (NHANES) estimated TBI prevalence to be 18.1% and 20.5% among foreign-born persons in 1999-2000 and 2011-2012 respectively.^25–27^ Other surveys in the United Kingdom and the Netherlands using interferon-gamma release assays estimated a TBI prevalence of approximately 20% among foreign-born persons between 2008 and 2012.^28,29^

The proportion of foreign-born residents in Canada from countries with TB disease incidence ≥50 per 100,000 persons increased to 55% by the 2016 census year, with a substantial increase in those from countries with incidence ≥200 per 100,000 persons. This was balanced by a decrease in estimated TBI prevalence among Canadians from some of the highest TB incidence countries, including China, India, and Viet Nam. This decline may potentially reflect the impact of both improved socioeconomic conditions and TB care programs in those countries. ^20,30^

These results can help inform clinical and programmatic decision-making around TBI screening and treatment. They provide a provincial-level picture of TBI prevalence, which is relevant as healthcare policies fall within the provincial purview. They also have implications for the development of TBI screening and treatment programs. Our results reinforce earlier suggestions that screening and providing TPT to people who immigrated before adulthood will have limited public health impact as TBI prevalence among this group is low.^13,31^ Even though TBI prevalence among older persons is higher, the risk of adverse events with TPT—even newer rifamycin-based regimens—increases with age^32,33^ and the likelihood the infection occurred within the previous 2 years is lower. Even if TBI testing targets persons from high-burden countries shortly after arrival, a small minority of persons found to have TBI will have been infected within the last two years. As the yield and cost-effectiveness of any TBI screening and treatment program increase with both pretest probability of TBI and risk of progression to TB in those with TBI,^11–13,28,31^ our results are directly relevant to future TB prevention programming.

Strengths of this analysis include the use of robust, previously reported methodology for estimating risks of acquiring TB infection over time in many countries. We also accounted explicitly for uncertainty around these country- and time-based estimates of infection risk.

### Limitations

Our analysis had several potential limitations. First, the source data assumed that individuals with positive TST results truly had had TBI and remained infected with viable mycobacteria for life, with no allowance for self-clearance. Hence we may have overestimated the prevalence of TBI, particularly in older age groups.^34,35,36^ However, this is also a potential shortcoming of any analysis based on current immunological tests for TBI. Indeed, it has been argued that failure to account for self-clearance leads to an underestimate of the number of people who have recently acquired TBI, and of their ensuing risk for TB disease and subsequent transmission.^37^ Second, we assumed that the ARI was consistent among persons within a given country in any given year, which obscures variation due to geography, mixing patterns, and other social factors.^38,39^ For example, refugees are more likely to have TBI and to progress to TB disease than other migrants from the same countries.^38^

Similarly, we lacked census data stratified by immigration category (e.g. refugee, economic class, family reunification) as well as sex-stratified ARI data, making us unable to evaluate TBI according to immigration category or sex.^39^ In tabulating TBI prevalence according to TB incidence in migrants’ countries of origin, we relied on recent incidence estimates rather than those corresponding to the dates they moved to Canada—as robust data were not consistently available before 2000. Hence some older foreign-born residents from countries with decreasing TB incidence faced higher past infection risks than current incidence estimates might imply. We did account for changing annual infection probabilities in our cumulative risk estimates. Finally, we could not estimate infection risks from 2016 to 2021, making us unable to evaluate any potential impact of the COVID-19 pandemic on TBI prevalence.

## CONCLUSION

Approximately one quarter of foreign-born Canadians likely had TBI from 2001 to 2016, but few were recently infected. The composition of the Canadian foreign-born population has changed, with more persons from countries with high TB disease incidence, although estimated prevalence has dropped somewhat among immigrants from several key countries. These data can help inform future strategies to reduce the burden of TB disease among foreign-born Canadians.

## Data Availability

All data produced in the present work are contained in the manuscript

https://github.com/Acesa2t

## REFERENCES

1 Houben RMGJ, Dodd PJ. The Global Burden of Latent Tuberculosis Infection: A Re-estimation Using Mathematical Modelling. PLOS Med 2016; 13: e1002152.

2 Gupta RK, Calderwood CJ, Yavlinsky A, et al. Discovery and validation of a personalized risk predictor for incident tuberculosis in low transmission settings. Nat Med 2020; 26: 1941–9.

3 World Health Organization [WHO]. Global tuberculosis report 2021. Geneva: World Health Organization, 2021 https://apps.who.int/iris/handle/10665/346387 (accessed Feb 5, 2022).

4 Canadian Tuberculosis Standards - 8th Edition. | Content Curation & Alerts in Medicine. LinksMedicus - Free Med. News Perspect. https://linksmedicus.com/ (accessed July 2, 2022).

5 Canada PHA of. Tuberculosis in Canada, 2020. 2022; published online March 23. https://www.canada.ca/en/public-health/services/publications/science-research-data/tuberculosis-canada-2020-infographic.html (accessed July 2, 2022).

6 Tuberculosis prevention and control in Canada: a federal framework for action. Ottawa, Ontario: Centre for Communicable Diseases and Infection Control, Public Health Agency of Canada, 2014.

7 Government of Canada SC. 2021 Census of Population - Data products. 2021; published online Nov 17. https://www12.statcan.gc.ca/census-recensement/2021/dp-pd/index-eng.cfm (accessed Nov 4, 2022).

8 BCCDC TB Annual Report 2018. http://www.bccdc.ca/resource-gallery/Documents/Statistics%20and%20Research/Statistics%20and%20Reports/TB/TB_Annual_Report_2018_website.pdf (accessed Jan 30, 2023).

9 Ronald LA, FitzGerald JM, Bartlett-Esquilant G, et al. Treatment with isoniazid or rifampin for latent tuberculosis infection: population-based study of hepatotoxicity, completion and costs. Eur Respir J 2020; 55: 1902048.

10 Campbell J, Marra F, Cook V, Johnston J. Screening immigrants for latent tuberculosis: Do we have the resources? CMAJ 2014; 186: 246–7.

11 Campbell JR, Johnston JC, Cook VJ, Sadatsafavi M, Elwood RK, Marra F. Cost-effectiveness of Latent Tuberculosis Infection Screening before Immigration to Low-Incidence Countries. Emerg Infect Dis 2019; 25: 661–71.

12 Campbell JR, Johnston JC, Sadatsafavi M, Cook VJ, Elwood RK, Marra F. Cost-effectiveness of post-landing latent tuberculosis infection control strategies in new migrants to Canada. PLOS ONE 2017; 12: e0186778.

13 Dale KD, Abayawardana MJ, McBryde ES, Trauer JM, Carvalho N. Modelling the Cost-Effectiveness of Latent Tuberculosis Screening and Treatment Strategies in Recent Migrants to a Low Incidence Setting. Am J Epidemiol 2021; published online May 21. DOI:10.1093/aje/kwab150.

14 Cauthen GM, Pio A, ten Dam HG. Annual risk of tuberculous infection. 1988. Bull World Health Organ 2002; 80: 503–11; discussion 501-502.

15 WHO. Global tuberculosis report 2015. World Health Organization, 2015 https://apps.who.int/iris/handle/10665/191102 (accessed Oct 9, 2022).

16 van Leth F. Prevalence of tuberculous infection and incidence of tuberculosis; a re-assessment of the Styblo rule. Bull World Health Organ 2008; 86: 20–6.

17 Trunz BB, Fine PEM, Dye C. Effect of BCG vaccination on childhood tuberculous meningitis and miliary tuberculosis worldwide: a meta-analysis and assessment of cost-effectiveness. The Lancet 2006; 367: 1173–80.

18 Dorfman R. A Note on the δ-Method for Finding Variance Formula. Biom Bull 1938; : 1: 129–137.

19 Government of Canada SC. Census Program: Previous censuses. 2001; published online Jan 15. https://www12.statcan.gc.ca/census-recensement/pc-eng.cfm (accessed Nov 4, 2022).

20 Global Tuberculosis Reports. https://www.who.int/teams/global-tuberculosis-programme/tb-reports (accessed Jan 31, 2023).

21 World Bank. “Incidence of tuberculosis (per 100,000 people)”. Indicators from World Bank: Data. https://data.worldbank.org/indicator/SH.TBS.INCD (accessed Jan 31, 2023).

22 WHO. WHO TB burden estimates. Geneva, World Health Organization. https://www.who.int/teams/global-tuberculosis-programme/data (accessed Jan 31, 2023).

23 WHO operational handbook on tuberculosis (Module 1 – Prevention): Tuberculosis preventive treatment. Geneva, World Health Organization. 2020. https://apps.who.int/iris/bitstream/handle/10665/331525/9789240002906-eng.pdf (accessed Jan 31, 2023).

24 Dale KD, Trauer JM, Dodd PJ, Houben Rmgj, Denholm JT. Estimating the prevalence of latent tuberculosis in a low-incidence setting: Australia. Eur Respir J 2018; 52: 1801218.

25 Miramontes R, Hill AN, Woodruff RSY, et al. Tuberculosis Infection in the United States: Prevalence Estimates from the National Health and Nutrition Examination Survey, 2011-2012. PLOS ONE 2015; 10: e0140881.

26 Grieco EM, Acosta YD, de la Cruz GP, et al. U.S. Census Bureau. American Community Survey Reports. Series ACS-19, The Foreign-Born Population in the United States: 2010. Retrieved from https://www.census.gov/library/publications/2012/acs/acs-19.html

27 Schmidley, A. Dianne, U.S. Census Bureau, Current Population Reports, Series P23-206, Profile of the Foreign-Born Population in the United States: 2000, U.S. Government Printing Office, Washington, DC, 2001.

28 Pareek M, Watson JP, Ormerod LP, et al. Screening of immigrants in the UK for imported latent tuberculosis: a multicentre cohort study and cost-effectiveness analysis. Lancet Infect Dis 2011; 11: 435–44.

29 Mulder C, Deutekom H van, Huisman EM, et al. Role of the QuantiFERON®-TB Gold In-Tube assay in screening new immigrants for tuberculosis infection. Eur Respir J 2012; 40: 1443–9.

30 Oxlade O, Schwartzman K, Behr MA, et al. Global tuberculosis trends: a reflection of changes in tuberculosis control or in population health? Int J Tuberc Lung Dis 2009; 13: 1238–46.

31 Shedrawy J, Deogan C, Öhd JN, et al. Cost-effectiveness of the latent tuberculosis screening program for migrants in Stockholm Region. Eur J Health Econ 2021; 22: 445–54.

32 Campbell JR, Trajman A, Cook VJ, et al. Adverse events in adults with latent tuberculosis infection receiving daily rifampicin or isoniazid: post-hoc safety analysis of two randomised controlled trials. Lancet Infect Dis 2020; 20: 318–29.

33 Pease C, Hutton B, Yazdi F, et al. A systematic review of adverse events of rifapentine and isoniazid compared to other treatments for latent tuberculosis infection. Pharmacoepidemiol Drug Saf 2018; 27: 557–66.

34 Behr MA, Edelstein PH, Ramakrishnan L. Is Mycobacterium tuberculosis infection life long? BMJ 2019; 367: 5770.

35 Emery JC, Richards AS, Dale KD, et al. Self-clearance of Mycobacterium tuberculosis infection: implications for lifetime risk and population at-risk of tuberculosis disease. Proc R Soc B Biol Sci 2021; 288: 20201635.

36 Opie EL. Pathology of the Tuberculosis of Childhood and Its Bearing on Clinical Work. Br Med J 1927; 2: 1130–5.

37 Dowdy DW, Behr MA. Are we underestimating the annual risk of infection with Mycobacterium tuberculosis in high-burden settings? Lancet Infect Dis 2022; 22: e271–8.

38 Styblo K. Epidemiology of Tuberculosis. Hague R Neth Tuberc Assoc 1991.

39 Ronald LA, Campbell JR, Balshaw RF, et al. Demographic predictors of active tuberculosis in people migrating to British Columbia, Canada: a retrospective cohort study. CMAJ 2018; 190: E209–16.

