## Appendix for "The Prevalence of Tuberculosis Infection Among Foreign-Born Canadians: A Modelling Study"

### Table S1. List of countries excluded from analysis

| **Excluded Countries of Origin** | |
| --- | --- |
| Aruba | Maldives |
| Anguilla | Marshall Islands |
| Andorra | Malta |
| Netherlands Antilles | Northern Mariana Islands |
| American Samoa | Montserrat |
| Antigua and Barbuda | New Caledonia |
| Bonaire, Sint Eustatius and Saba | Niue |
| Bahamas | Nauru |
| Belize | Palau |
| Bermuda | French Polynesia |
| Barbados | Samoa |
| Brunei Darussalam | San Marino |
| Cook Islands | Sao Tome and Principe |
| Curacao | Sint Maarten (Dutch part) |
| Cayman Islands | Seychelles |
| Dominica | Turks and Caicos Islands |
| Micronesia, Federated States of | Tokelau |
| Grenada | Timor-Leste |
| Greenland | Tonga |
| Guam | Tuvalu |
| Iceland | Saint Vincent and the Grenadines |
| Kiribati | Virgin Islands, British |
| Saint Kitts and Nevis | U.S. Virgin Islands |
| Saint Lucia | Vanuatu |
| Monaco | Wallis and Futuna |

### Table S2. Table of regions, countries, and special administrative regions included in the analysis

| **Regions** | **Countries** | **Special Administrative Regions / Jurisdictions** |
| --- | --- | --- |
| Americas; Europe; Africa; Asia; Oceania | Afghanistan; Albania; Algeria; American Samoa; Andorra; Angola; Anguilla; Antigua and Barbuda; Argentina; Armenia; Aruba; Australia; Austria; Azerbaijan; Bahamas; Bahrain; Bangladesh; Barbados; Belarus; Belgium; Belize; Benin; Bermuda; Bhutan; Bolivia; Bosnia and Herzegovina; Botswana; Brazil; Brunei Darussalam; Bulgaria; Burkina Faso; Burma (Myanmar); Burundi; Cabo Verde; Cambodia; Cameroon; Cayman Islands; Central African Republic; Chad; Chile; China; Colombia; Comoros; Congo, Democratic Republic of the; Congo, Republic of the; Cook Islands; Costa Rica; Croatia; Cuba; Cùte d’Ivoire; Cyprus; Czech Republic; Denmark; Djibouti; Dominica; Dominican Republic; Ecuador; Egypt; El Salvador; Equatorial Guinea; Eritrea; Estonia; Ethiopia; Federated States of Micronesia; Fiji; Finland; France; French Polynesia; Gabon; Gambia; Georgia; Germany; Ghana; Greece; Greenland; Grenada; Guam; Guatemala; Guinea; Guinea-Bissau; Guyana; Haiti; Honduras; Hong Kong; Hungary; Iceland; India; Indonesia; Iran; Iraq; Ireland; Israel; Italy; Jamaica; Japan; Jordan; Kazakhstan; Kenya; Kiribati; Korea, North; Korea, South; Kuwait; Kyrgyzstan; Laos; Latvia; Lebanon; Lesotho; Liberia; Libya; Lithuania; Luxembourg; Macau; Macedonia, Republic of; Madagascar; Malawi; Malaysia; Maldives; Mali; Malta; Marshall Islands; Mauritania; Mauritius; Mexico; Moldova; Monaco; Mongolia; Montenegro; Montserrat; Morocco; Mozambique; Namibia; Nepal; Netherlands; New Caledonia; New Zealand; Nicaragua; Niger; Nigeria; Northern Mariana Islands; Norway; Oman; Pakistan; Palestine; Panama; Papua New Guinea; Paraguay; Peru; Philippines; Poland; Portugal; Puerto Rico; Qatar; Romania; Russian Federation; Rwanda; Saint Kitts and Nevis; Saint Lucia; Saint Vincent and the Grenadines; Samoa; Sao Tome and Principe; Saudi Arabia; Senegal; Serbia; Seychelles; Sierra Leone; Singapore; Slovakia; Slovenia; Somalia; South Africa, Republic of; Spain; Sri Lanka; Sudan; Suriname; Swaziland; Sweden; Switzerland; Syria; Tajikistan; Tanzania; Thailand; Timor-Leste; Togo; Tonga; Trinidad and Tobago; Tunisia; Turkey; Turkmenistan; Uganda; Ukraine; United Kingdom; United States; Uruguay United Arab Emirates; Uzbekistan; Vanuatu; Venezuela; Viet Nam; Virgin Islands, British; Wallis and Futuna; Yemen; Zambia; Zimbabwe | Hong Kong  Macau |

### Table S3. Initial census data variables as provided by Statistics Canada.

| **Age Groups** | | | |
| --- | --- | --- | --- |
| 0 to 14 years | | | |
| 15 to 24 years | | | |
| 25 to 54 years | | | |
| 55 to 64 years | | | |
| 65 years and older | | | |
| **Place of Birth** | | | |
| Regions* | | | |
| Sub-Regions** | | | |
| Countries | | | |
| Special Administrative Regions | | | |
| **Year of Immigration** | | | |
| **Census 2001†** | **Census 2006†** | **Census 2011** | **Census 2016** |
| Before 1915 | Same as Census 2001 | Before 1971 | Before 1981 |
| 1916 to 1920 | 2001 to 2006 | 1971 to 1980 | 1981 to 1990 |
| 1921 to 1925 |  | 1981 to 1990 | 1991 to 2000 |
| 1926 to 1930 |  | 1991 to 2000 | 2001 to 2010 |
| 1931 to 1935 |  | 2001 to 2011 | 2001 to 2005 |
| 1936 to 1940 |  |  | 2006 to 2010 |
| 1941 to 1945 |  |  | 2011 to 2016 |
| 1946 to 1950 |  |  |  |
| 1951 to 1955 |  |  |  |
| 1956 to 1960 |  |  |  |
| 1961 to 1965 |  |  |  |
| 1966 to 1970 |  |  |  |
| 1970 to 1975 |  |  |  |
| 1976 to 1980 |  |  |  |
| 1981 to 1985 |  |  |  |
| 1986 to 1990 |  |  |  |
| 1991 to 1995 |  |  |  |
| 1996 to 2001 |  |  |  |

**Regions are defined as Americas, Europe, Africa, Asia, and Oceania.*

***Special Administrative Regions included Hong Kong and Macau.*

*†The 2001 and 2006 census data were provided by Statistics Canada's Statistical Consultation Group*

### Table S4. Estimated tuberculosis infection prevalence by age at immigration and TB disease incidence in country of origin per 100,000 persons during census years 2001, 2006, 2011 and 2016.

| TB Infection Prevalence in 2001 (%, 95% UI) | | | | | | |
| --- | --- | --- | --- | --- | --- | --- |
|  | Age at Immigration <15 | Age at Immigration 15-34 | Age at Immigration 35-54 | Age at Immigration 55-74 | Age at Immigration ≥75 | All |
| TB Incidence in Country of Origin per 100,000 persons |  |  |  |  |  |  |
| 0-9 | 6 (2-24) | 15 (7-36) | 21 (11-44) | 25 (10-53) | 31 (12-61) | 13 (6-31) |
| 10-49 | 7 (7-3-18) | 22 (12-41) | 33 (20-50) | 45 (27-64) | 49 (31-69) | 21 (12-36) |
| 50-99 | 8 (5-16) | 28 (16-42) | 48 (28-68) | 71 (37-87) | 75 (37-91) | 32 (19-44) |
| 100-199 | 15 (8-32) | 50 (25-73) | 69 (37-86) | 83 (37-97) | 86 (47-97) | 46 (23-64) |
| 200+ | 13 (10-19) | 40 (29-54) | 58 (42-72) | 72 (52-89) | 79 (58-94) | 40 (29-52) |
| All | 8 (6-18) | 25 (19-37) | 41 (33-50) | 59 (45-70) | 66 (51-76) | 25 (20-35) |
| TB Infection Prevalence in 2006 (%, 95% UI) | | | | | | |
|  | Age at Immigration <15 | Age at Immigration 15-34 | Age at Immigration 35-54 | Age at Immigration 55-74 | Age at Immigration ≥75 | All |
| TB Incidence in Country of Origin per 100,000 persons |  |  |  |  |  |  |
| 0-9 | 6 (2-23) | 15 (7-34) | 18 (10-38) | 23 (10-48 | 30 (12-60) | 12 (6-28) |
| 10-49 | 6 (3-15) | 20 (11-35) | 30 (19-44) | 42 (27-60) | 48 (31-65) | 18 (11-31) |
| 50-99 | 7 (4-12) | 26 (15-38) | 46 (28-64) | 69 (36-86) | 73 (42-89) | 30 (18-41) |
| 100-199 | 14 (8-29) | 46 (24-66) | 63 (38-80) | 82 (42-95) | 83 (55-95) | 42 (23-58) |
| 200+ | 12 (10-16) | 38 (29-50) | 57 (43-69) | 71 (54-88) | 79 (62-93) | 39 (30-49) |
| All | 8 (6-16) | 25 (19-34) | 40 (33-49) | 59 (47-69) | 67 (54-75) | 24 (20-33) |
| TB Infection Prevalence in 2011 (%, 95% UI) | | | | | | |
|  | Age at Immigration <15 | Age at Immigration 15-34 | Age at Immigration 35-54 | Age at Immigration 55-74 | Age at Immigration ≥75 | All |
| TB Incidence in Country of Origin per 100,000 persons |  |  |  |  |  |  |
| 0-9 | 7 (3-26) | 12 (6-27) | 15 (7-29) | 20 (8-41) | 25 (10-52) | 10 (5-27) |
| 10-49 | 7 (4-17) | 16 (10-26) | 25 (17-35) | 38 (25-55) | 42 (27-62) | 16 (10-25) |
| 50-99 | 7 (4-12) | 22 (14-31) | 42 (26-56) | 63 (35-82) | 71 (41-88) | 26 (16-35) |
| 100-199 | 14 (8-26) | 39 (24-56) | 57 (38-72) | 78 (44-92) | 81 (57-95) | 37 (23-51) |
| 200+ | 12 (10-15) | 36 (29-46) | 56 (45-68) | 69 (54-84) | 78 (63-92) | 37 (30-46) |
| All | 8 (6-18) | 23 (19-29) | 39 (33-46) | 57 (46-66) | 64 (52-73) | 23 (19-30) |
| TB Infection Prevalence in 2016 (%, 95% UI) | | | | | | |
|  | Age at Immigration <15 | Age at Immigration 15-34 | Age at Immigration 35-54 | Age at Immigration 55-74 | Age at Immigration ≥75 | All |
| TB Incidence in Country of Origin per 100,000 persons |  |  |  |  |  |  |
| 0-9 | 6 (2-21) | 11 (5-22) | 13 (6-23) | 17 (8-35) | 22 (9-47) | 9 (5-22) |
| 10-49 | 7 (4-15) | 14 (9-24) | 22 (15-31) | 34 (23-52) | 39 (24-61) | 14 (9-22) |
| 50-99 | 7 (4-13) | 21 (14-29) | 39 (25-51) | 60 (34-79) | 72 (38-90) | 24 (16-33) |
| 100-199 | 14 (7-24) | 36 (23-50) | 53 (38-66) | 75 (50-89) | 78 (58-93) | 34 (22-46) |
| 200+ | 12 (9-15) | 35 (29-42) | 54 (44-64) | 67 (54-83) | 75 (62-90) | 35 (22-46) |
| All | 8 (6-16) | 22 (19-26) | 37 (31-43) | 55 (45-63) | 65 (50-74) | 22 (19-28) |

*UI = uncertainty interval*

Between census years 2001 and 2016, TBI prevalence by age at immigration did not change substantially. Prevalence decreased by TB disease in country of origin for all groups, but most notably among foreign-born Canadians from countries with TB disease incidence of 100-199 per 100,000 persons from 46% in 2001 to 34% in 2016. Additionally, for census years 2001 and 2006, prevalence among persons from countries with incidence of 100-199 per 100,000 persons remained higher than that of those from countries with incidence ≥200 per 100,000; however, the difference in prevalence decreased between each census year (Table S4).

### Table S5. Numbers and ages of foreign-born Canadians in provinces of Alberta, British Columbia, Ontario, Quebec, and all other provinces and territories in 2016.

| **Province of Residence** | **Foreign-Born Residents (N)** | **Median Age in 2016 (IQR)** | **Median Age at Immigration (IQR)** |
| --- | --- | --- | --- |
| Alberta | 845,215 | 44 (30-57) | 23 (10-35) |
| British Columbia | 1,292,675 | 50 (35-65) | 23 (10-36) |
| Ontario | 3,852,150 | 49 (35-64) | 22 (10-34) |
| Quebec | 1,091,305 | 44 (30-57) | 23 (10-35) |
| Other* | 459,750 | 44 (29-59) | 24 (8-43) |
| **All** | 7,539,895 | 48 (33-62) | 23 (10-35) |

*N = number, IQR = interquartile range*

**Other is defined as Nova Scotia, New Brunswick, Manitoba, Prince Edward Island, Saskatchewan, Newfoundland and Labrador, Northwest Territories, Yukon, and Nunavut.*

### Table S6. Estimated tuberculosis infection prevalence by age in census year and years since immigration

| **Tuberculosis Infection Prevalence in 2016 (%, 95% UI)** | | | | | | |
| --- | --- | --- | --- | --- | --- | --- |
| **Years Since**  **Immigration** | **Age in Census Year 2016** | | | | | |
|  | **0-14** | **15-34** | **35-54** | **55-74** | **75+** | **All** |
| **0-4** | 3 (3-3) | 16 (15-17) | 32 (28-36) | 49 (40-59) | 65 (49-75) | 23 (21-25) |
| **5-9** | 2 (1-2) | 12 (11-13) | 29 (26-33) | 46 (39-55) | 61 (49-70) | 22 (20-25) |
| **10-14** | 1 (1-1) | 9 (9-10) | 27 (24-32) | 45 (37-53) | 59 (48-68) | 22 (19-25) |
| **15-19** | *N/A* | 6 (6-7) | 24 (20-28) | 43 (34-52) | 58 (47-68) | 24 (20-28) |
| **20+** | *N/A* | 4 (4-5) | 14 (11-17) | 25 (19-35) | 33 (23-51) | 22 (17-30) |
| **All** | 2 (2-2) | 10 (9-10) | 23 (19-26) | 29 (24-39) | 37 (27-54) | 22 (20-28) |

*UI = Uncertainty Interval*

### Figure S1. Data structure and corresponding equations.

Equation 1 shows estimation of cumulative risk of infection (ROI) among foreign-born persons immigrating to Canada. Subscripts b, o, m, c, and f refer to birth year, country of origin, migration year, Canada, and final census year of calculation, respectively.

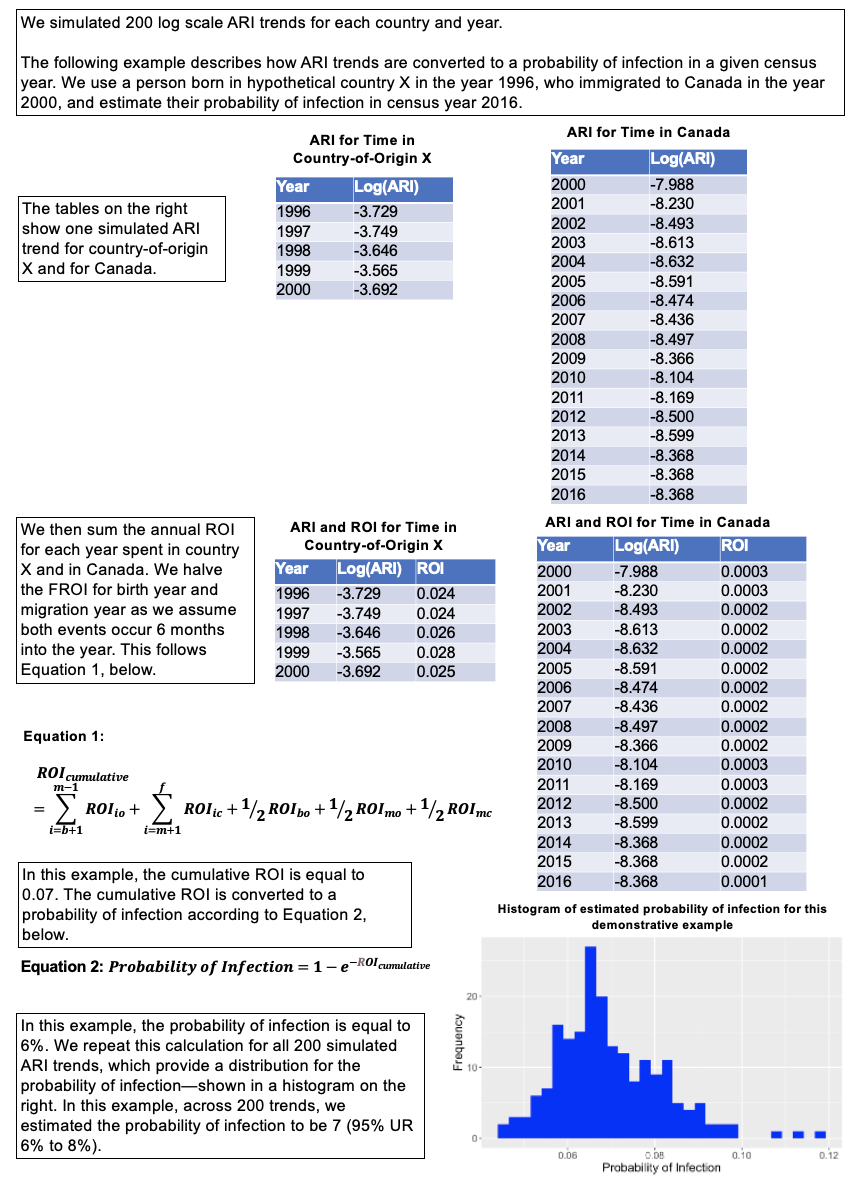
*Equation 2 shows how probability of infection among foreign-born persons was estimated using the cumulative ROI.*
